# Do maternal intrauterine environmental influences that lower offspring birthweight causally increase offspring cardiometabolic risk factors in later life? A Mendelian randomization study of 45,849 genotyped parent offspring pairs in the HUNT study

**DOI:** 10.1101/2020.05.04.20091173

**Authors:** Gunn-Helen Moen, Ben Brumpton, Cristen Willer, Bjørn Olav Åsvold, Kåre Birkeland, Michael C Neale, Rachel M Freathy, George Davey Smith, Deborah A Lawlor, Robert M Kirkpatrick, Nicole M Warrington, David M Evans

## Abstract

**Introduction:** There is a robust and well-documented observational relationship between lower birthweight and higher risk of cardiometabolic disease in later life. The Developmental Origins of Health and Disease (DOHaD) hypothesis posits that adverse environmental factors *in utero* or in the early years of life result in increased future risk of cardiometabolic disease. Our aim was to investigate whether there was evidence for causal effects of the intrauterine environment, as proxied by maternal single nucleotide polymorphisms (SNPs) that influence offspring birthweight independent of offspring genotype, on offspring cardiometabolic risk factors such as blood pressure, non-fasting glucose, body mass index (BMI), and lipid levels.

**Methods:** We investigated whether a genetic risk score of maternal SNPs associated with offspring birthweight was also associated with offspring cardiometabolic risk factors, after controlling for offspring genotypes at the same loci, in up to 26,057 mother-offspring pairs from the Nord-Trøndelag Health (HUNT) Study. We also conducted similar analyses in 19,792 father-offspring pairs from the same study to investigate whether there was evidence that any such causal effects operated through the postnatal, rather than the intrauterine environment. To take account of the considerable cryptic relatedness in HUNT, we implemented a computationally efficient genetic linear mixed model using the OpenMx software package to perform our analyses.

**Results:** We found little evidence for a maternal genetic effect of birthweight associated variants on offspring cardiometabolic risk factors after adjusting for offspring genotypes at the same loci. Likewise, we found little evidence for paternal genetic effects on offspring cardiometabolic risk factors performing similar analyses in father-offspring pairs. In contrast, offspring genetic risk scores of birthweight associated variants were strongly related to many cardiometabolic risk factors, even after conditioning on maternal genotypes at the same loci.

**Conclusion:** Our results suggest that the maternal intrauterine environment, as proxied by maternal SNPs that influence offspring birthweight, is unlikely to be a major determinant of adverse cardiometabolic outcomes in population based samples of individuals. In contrast, genetic pleiotropy appears to explain some of the observational relationship between offspring birthweight and future cardiometabolic risk.

## Introduction

There is a robust and well-documented observational relationship between lower birthweight and higher risk of cardiometabolic diseases in later life, including cardiovascular disease (CVD) and type 2 diabetes (T2D). The Developmental Origins of Health and Disease (DOHaD) hypothesis posits that adverse environmental factors *in utero* or in the early years of life result in increased future risk of cardiometabolic disease [1-8]. Evidence in favour of DOHaD has primarily come from observational [1-3] and animal studies [9], however, definitive causal evidence from human studies is lacking.

Mendelian randomization (MR) is an epidemiological method used to investigate whether an observational association between an exposure and an outcome represents a causal relationship [10]. Several studies have recently attempted to use MR to investigate the relationship between lower birthweight and cardiometabolic disease to inform on the validity of DOHaD [11-13]. However, these MR studies have used sub-optimal methodologies in which only offspring genotypes are considered as genetic instruments to proxy offspring birthweight [14]. This limitation contrasts strikingly with the argument that many DOHaD proponents would make i.e. that an adverse maternal environment during pregnancy, results in low birthweight and increased risk of future cardiometabolic disease [1, 5, 7]. This hypothesis is entirely distinct from postulating that birthweight itself has a direct causal effect on risk of cardiometabolic disease [14]. Thus, these early MR studies have ignored the potential contribution of the maternal genome (correlated 0.5 with the offspring genome[15, 16]), meaning that any association between offspring SNPs and offspring cardiometabolic risk may in fact be due to maternal genotypes, violating core assumptions underlying MR [17], and complicating interpretation of the results. Indeed, Davey Smith and Ebrahim (2003) in their initial description of the MR methodology, noted that the appropriate way of using MR to investigate the effects of the intrauterine environment on offspring outcomes (in their example maternal folate intake and offspring neural tube defects), was to use maternal genotypes to proxy the intrauterine environment [10].

We have previously described how MR principles can be harnessed to test aspects of DOHaD using maternal SNPs that are related to offspring birthweight and/or adverse maternal environmental exposures during pregnancy [14, 16, 18-20]. For example, one possibility is to test whether SNPs in the mother that are directly related to offspring birthweight are also associated with offspring cardiometabolic risk factors, after conditioning on offspring genotypes at the same loci. To understand why this analysis would be informative, consider Figure 1, which illustrates four credible ways in which maternal SNPs can simultaneously be related to offspring birthweight and future offspring cardiometabolic risk factors. In panel (a), maternal birthweight associated SNPs produce an *in utero* environment that leads to reduced fetal growth and subsequently low offspring birthweight and developmental compensations that produce increased risk of offspring cardiometabolic disease in later life. In panel (b), low offspring birthweight itself is causal for increased risk of offspring cardiometabolic disease. Under panels (a) and (b), the existence of a relationship between maternal alleles asociated with lower birthweight and higher cardiometabolic risk in the offspring (after conditioning on offspring genotype at the same loci) argues strongly in favour of a DOHaD mechanism, where developmental compensations to reduced fetal growth impact on future health. In panel (c), the inverse genetic correlation between offspring birthweight and offspring cardiometabolic disease is driven entirely by genetic pleiotropy in the offspring genome, and importantly, not via DOHaD mechanisms. Under this model, maternal genotypes related to lower offspring birthweight will not be associated with increased offspring cardiometabolic risk after conditioning on offspring genotype. Finally, in panel (d), SNPs that exert maternal effects on offspring birthweight also pleiotropically influence offspring cardiometabolic disease through the postnatal environment. If genotyped father-offspring pairs are also available, then paternal SNPs at the same loci can be tested for association with offspring cardiometabolic risk factors (conditional on offspring genotype). The existence of such associations would suggest that the post-natal environment (i.e. early life DOHaD influences such as via genetic nurture or dynastic effects rather than the intrauterine environment) may be responsible for the correlation between maternal genotypes and offspring cardiometabolic risk factors.

**Figure 1:**
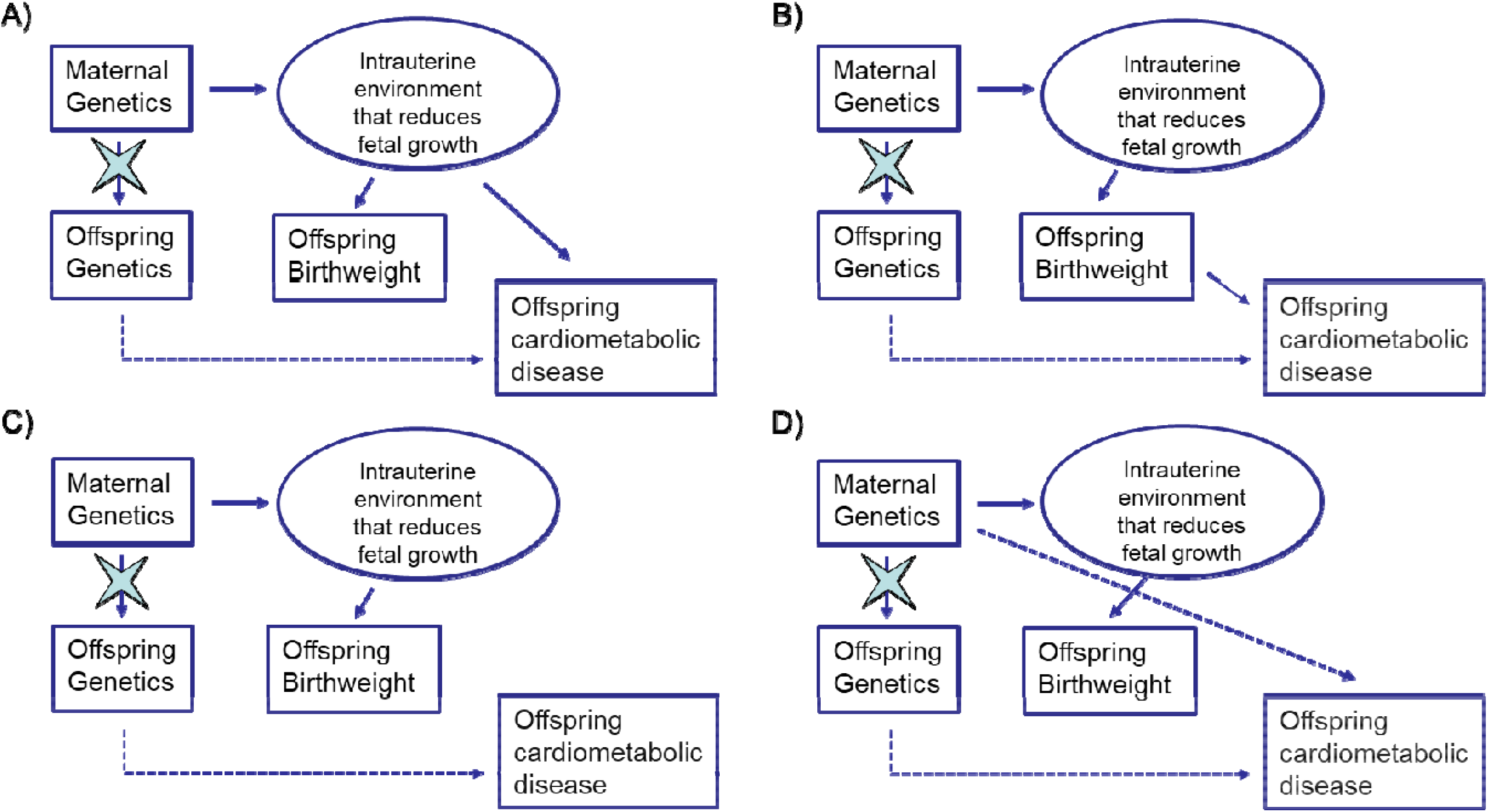
Four credible ways in which maternal SNPs can be related to offspring birthweight and offspring cardiometabolic risk factors: a) Maternal SNPs produce an adverse *in utero* environment that leads to fetal growth restriction and subsequently low offspring birthweight and developmental compensations that produce increased risk of offspring cardiometabolic disease in later life. b) Maternal SNPs produce an adverse *in utero* environment that leads to fetal growth restriction and low offspring birthweight. Low offspring birthweight in turn is causal for increased risk of offspring cardiometabolic disease. c) Maternal SNPs produce an adverse *in utero* environment that leads to fetal growth restriction and reduced birthweight. The same SNPs are transmitted to the offspring and pleiotropically influence offspring cardiometabolic risk through the offspring genome. d) Maternal SNPs produce an adverse *in utero* environment that leads to fetal growth restriction and reduced offspring birthweight. SNPs that exert maternal effects on offspring birthweight also pleiotropically influence offspring cardiometabolic disease through the postnatal environment. The star on the arrows denotes the act of conditioning on maternal or offspring genotype blocking the association between maternal and offspring variables. The dotted paths indicate paths in which the maternal genotype can be related to offspring phenotype that are not to do with intrauterine growth restriction. Finally, we note that some offspring SNPs may also exert direct effects on offspring birthweight (these not shown). The presence of direct effects from offspring genotype on offspring birthweight is inconsequential so long as the relevant analyses are conditional on offspring genotype.

In other words, the presence of correlation between maternal genotypes and offspring cardiometabolic risk factors, after conditioning on offspring genotypes at the same loci, is highly suggestive of DOHaD mechanisms related to lower birth weight (providing these associations are not replicated in father-offspring pairs also). We emphasize that the paradigm illustrated in Figure 1, which we use in our study, only tests one aspect of DOHaD (i.e. that maternal exposures that affect offspring birthweight are also causal for increased offspring cardiometabolic risk). It is possible that there are other maternal exposures that affect the offspring prenatal or postnatal environment, but do not influence offspring birthweight, and still affect future offspring cardiometabolic risk. We do not test for the influence of these exposures on offspring cardiometabolic risk in this study, but limit our attention to those that exert an effect on offspring birthweight (a distinction we explore further in the discussion).

We have previously used this paradigm to examine the association between maternal birthweight related SNPs and offspring blood pressure in the UK Biobank study as a preliminary test of the validity of this possible DOHaD mechanism [18]. Interestingly, this showed that maternal SNPs related to low offspring birthweight, were actually associated with *lower* offspring systolic blood pressure after conditioning on offspring genotype at the same loci (i.e. the *opposite* of what would be expected if maternal intrauterine effects that reduce fetal growth result in higher later-life cardiometabolic risk). However, the number of mother offspring pairs used in this previous study was small (N = 3,886) and systolic blood pressure was the only cardiometabolic risk factor investigated. Therefore, the results from this preliminary study need to be replicated and further cardiometabolic risk factors examined. The Norwegian based HUNT Study [21], which contains approximately 70,000 genotyped individuals, including 45,849 parent-offspring pairs, is one of the few cohorts where such analyses can be conducted. The average age of the HUNT offspring is approximately 40 years, rendering this cohort not only one of the largest cohorts in the world with genotyped mother-offspring pairs (and father offspring pairs) with birthweight information, but also one of the few with offspring old enough to have developed adverse cardiometabolic profiles.

In the present manuscript, we performed genetic association analyses in up to 26,057 genotyped mother-offspring pairs from the Norwegian HUNT Study in order to investigate whether there was evidence for a causal effect of the intrauterine environment (proxied by maternal SNPs that influence offspring birthweight) on offspring cardiometabolic risk factors. We investigated whether maternal genotypes associated with lower offspring birthweight were also associated with later life offspring cardiometabolic risk factors such as blood pressure, non-fasting glucose levels, body mass index (BMI), and lipid levels, after conditioning on offspring genotype at the same loci. We also performed similar analyses in up to 19,792 father-offspring pairs to investigate whether there was evidence for a post-natal environmental effect (genetic nurture or dynastic effects), rather than an intrauterine environmental effect. In the course of executing these analyses, we implement a computationally efficient genetic linear mixed model that not only enables the investigation of causal questions relevant to the specific DOHaD mechanism that is the focus of this paper, but also simultaneously accounts for the considerable cryptic relatedness within the HUNT Study.

## Methods

### HUNT Study

The Nord-Trøndelag Health Study (HUNT) is a large population-based health study of the inhabitants of Nord-Trøndelag County in central Norway that commenced in 1984. A comprehensive description of the study population has been previously reported [21]. Approximately every 10 years the entire adult population of Nord-Trøndelag (∼90,000 adults in 1995) is invited to attend a health survey which includes comprehensive questionnaires, an interview, clinical examination, and detailed phenotypic measurements (HUNT1 (1984 to 1986); HUNT2 (1995 to 1997); HUNT3 (2006 to 2008) and HUNT4 (2017 to 2019)). These surveys have high participation, with 89%, 69%, 54% and 54% of invited adults participating in HUNT1, 2, 3 and 4, respectively [21, 22]. Additional phenotypic information is collected by integrating national registers. Approximately 90% of participants from HUNT2 and HUNT3 were genotyped in 2015 [23].

The HUNT Study was approved by the Regional Committee for Medical and Health Research Ethics, Norway and all participants gave informed written consent (REK Central application number 2018/2488).

### Genotyping, quality control and imputation

Genotyping, quality control and imputation in the HUNT cohort have been described in detail elsewhere [23]. In short, DNA from 71,860 HUNT samples were genotyped using one of three different Illumina HumanCoreExome arrays (HumanCoreExome12 v1.0, HumanCoreExome12 v1.1 and UM HUNT Biobank v1.0). Genomic position, strand orientation and the reference allele of genotyped variants were determined by aligning their probe sequences against the human genome (Genome Reference Consortium Human genome build 37 and revised Cambridge Reference Sequence of the human mitochondrial DNA; http://genome.ucsc.edu) using BLAT [24]. Ancestry of all samples was inferred by projecting all genotyped samples into the space of the principal components of the Human Genome Diversity Project (HGDP) reference panel (938 unrelated individuals; downloaded from http://csg.sph.umich.edu/chaolong/LASER/) [25, 26], using PLINK v1.90 [27]. The resulting genotype data were phased using Eagle2 v2.3 [28]. Imputation was performed on the 69,716 samples of recent European ancestry using Minimac3 (v2.0.1, http://genome.sph.umich.edu/wiki/Minimac3) [29] with default settings (2.5 Mb reference based chunking with 500kb windows) and a customized Haplotype Reference consortium release 1.1 (HRC v1.1) for autosomal variants and HRC v1.1 for chromosome X variants [30].

### Identifying genotyped parent-offspring pairs

Before the kinship analysis, the plink files with genotyped SNPs underwent a second stage of cleaning. Any individuals whose inferred sex contradicted their reported gender (N=348) as well as individuals showing high or low heterozygosity (+/- 5SD from the mean) (N=412) were removed (760 individuals in total). In addition, variants with minor allele frequency <0.005 or more than 5% missing rate were removed. Parent-offspring pairs were identified by kinship analysis using the KING software[31]. Only genotyped SNPs shared across the arrays on autosomal chromosomes were used for the analysis – a total of 257,488 SNPs.

From the analysis, 46,428 parent-offspring relationships were identified, in addition to 35,373 full siblings, 128,334 second degree relationships and 386,619 third degree relationships based on the kinship analysis performed using the KING software and recommended thresholds for relatedness implemented as part of this package [31]. Any parent-offspring pair with 15 years or fewer difference in birth year was removed from further analyses. After removing these pairs, a total of 26,057 mother-offspring pairs and 19,792 father-offspring pairs of European ancestry with genotype information passing QC were identified. Each parent had between one and eight offspring available for analysis. Supplementary Table 1 shows the number of offspring per mother/father available for analysis.

**Table 1:** Descriptive statistics for offspring cardiometabolic risk factors in the phenotypic association analyses

| Phenotype | Mother-Offspring pairs |  |  |  | Father-Offspring pairs |  |  |  |
| --- | --- | --- | --- | --- | --- | --- | --- | --- |
|  | N | Mean | SD | Range | N | Mean | SD | Range |
| Birthweight (g) | 7,825 | 3570 | 482.2 | 1390-5900 | 6,875 | 3572 | 480.2 | 1660-5900 |
| Age | 7,825 | 30.1 | 6.6 | 19.1-41.4 | 6,875 | 30.1 | 6.6 | 19.2-41.4 |
| Sex (% male) | 7,825 | 45.3 | - | - | 6,875 | 45.7 | - | - |
| SBP (mmHg) | 7,792 | 122.9 | 13.4 | 70.0-207.0 | 6,846 | 122.8 | 13.4 | 70.0-207.0 |
| DBP (mmHg) | 7,790 | 69.0 | 9.8 | 36.9-117.0 | 6,845 | 68.9 | 9.7 | 38.0-120.0 |
| Glucose (mmol/L) | 7,659 | 4.95 | 1.16 | 2.29-11.95 | 6,727 | 4.95 | 1.17 | 2.29-11.47 |
| Total Cholesterol (mmol/L) | 7,684 | 4.89 | 0.97 | 2.00-9.90 | 6,749 | 4.88 | 0.97 | 2.30-9.90 |
| LDL Cholesterol (mmol/L) | 7,674 | 2.92 | 0.84 | 0.27-6.98 | 6,742 | 2.91 | 0.84 | 0.27-7.27 |
| HDL Cholesterol (mmol/L) | 7,682 | 1.31 | 0.32 | 0.50-2.80 | 6,748 | 1.32 | 0.32 | 0.50-2.80 |
| Triglycerides (mmol/L) | 7,786 | 1.22 | 1.72 | 0.30-11.25 | 6,839 | 1.21 | 1.72 | 0.30-11.25 |
| BMI | 7,803 | 25.79 | 1.19 | 15.64-49.40 | 6,853 | 25.28 | 1.17 | 15.96-49.40 |

SBP: systolic blood pressure; DBP: diastolic blood pressure; Glucose: non-fasting glucose; BMI: body mass index; LDL: low density lipoprotein; HDL: high density lipoprotein.

### Genetic risk scores

SNPs previously associated with own or offspring birthweight at genome-wide levels of significance in the Early Growth Genetics (EGG) Consortium paper [18] were extracted from the HUNT imputed genotype data in dosage format using plink2 [27]. Dosages were coded so that increasing dosages reflected maternal alleles associated with increased offspring birthweight based on conditional genome-wide association study (GWAS) results previously published [18]. Unweighted genetic risk scores (GRS) were constructed by simply adding the expected number of increasing birthweight alleles together for each individual. We used unweighted scores, because we do not know the extent to which each allele influences growth restriction, and so it does not make sense to weight scores by e.g. their observed effect on birthweight. Three GRS were constructed – one using all autosomal SNPs shown to have an effect on birthweight (N=204) from the recent EGG Consortium GWAS paper of birthweight [18] that found 205 autosomal SNPs, but rs9267812 was not available in the HUNT data), one using SNPs shown to have a maternal effect (N=71; i.e. some of these SNPs also had a fetal effect on birthweight), and one using SNPs that *only* had a significant maternal effect (N=31) (Supplementary Table 2).

**Table 2:** Association between offspring birthweight, offspring birthweight squared and offspring cardiometabolic risk factor in mother-offspring and father-offspring pairs.

| Phenotype | Mother-Offspring pairs |  |  |  |  |  |  | Father-Offspring pairs |  |  |  |  |  |  |
| --- | --- | --- | --- | --- | --- | --- | --- | --- | --- | --- | --- | --- | --- | --- |
|  | N | Birthweight |  |  | Birthweight Squared |  |  | N | Birthweight |  |  | Birthweight Squared |  |  |
|  |  | Effect size | SE | p-value | Effect size | SE | p-value |  | Effect size | SE | p-value | Effect size | SE | p-value |
| SBP (mmHg) | 7,792 | -3.71 | 2.60 | 0.15 | 0.35 | 0.36 | 0.33 | 6,846 | -4.51 | 2.85 | 0.11 | 0.43 | 0.39 | 0.27 |
| DBP (mmHg) | 7,790 | -1.95 | 2.01 | 0.33 | 0.16 | 0.28 | 0.56 | 6,845 | -3.14 | 2.13 | 0.14 | 0.32 | 0.30 | 0.29 |
| Glucose (mmol/L)* | 7,659 | **-0.08** | **0.04** | **0.02** | **0.01** | **4.8x10<sup>-3</sup>** | **0.04** | 6,727 | **-0.07** | **0.04** | **0.05** | **8.9x10<sup>-3</sup>** | **5.2x10<sup>-3</sup>** | 0.09 |
| Total Cholesterol (mmol/L) | 7,684 | -0.29 | 0.21 | 0.17 | 0.03 | 0.029 | 0.28 | 6,749 | -0.20 | 0.22 | 0.36 | 0.02 | 0.03 | 0.54 |
| LDL Cholesterol (mmol/L) | 7,674 | -0.32 | 0.18 | 0.08 | 0.04 | 0.03 | 0.11 | 7,674 | -0.25 | 0.19 | 0.18 | 0.03 | 0.03 | 0.23 |
| HDL Cholesterol (mmol/L) | 7,682 | **0.17** | **0.07** | **0.01** | **-0.02** | **0.01** | **0.02** | 6,748 | **0.16** | **0.07** | **0.03** | **-0.02** | **0.01** | **0.03** |
| Triglycerides (mmol/L)* | 7,786 | 0.03 | 0.03 | 0.28 | 0.02 | 0.02 | 0.19 | 6,839 | -0.22 | 0.12 | 0.07 | 0.02 | 0.02 | 0.16 |
| BMI* | 7,803 | -0.06 | 0.04 | 0.08 | **0.01** | **0.01** | **0.03** | 6,853 | -0.06 | 0.04 | 0.09 | **0.01** | **0.01** | **0.04** |

\*Offspring phenotype first (natural) logarithm transformed. SBP: systolic blood pressure; DBP: diastolic blood pressure; Glucose: non-fasting glucose; BMI: body mass index; LDL: low density lipoprotein; HDL: high density lipoprotein. All analyses are adjusted for age, sex and participation period. Results with p-values less than 0.05 are shown in bold.

### Outcome variables

During the health surveys (HUNT1-4) [21] clinical examination, and detailed phenotypic measurements were performed on all participants. For all cardiometabolic risk factors in the offspring (BMI, systolic blood pressure (SBP), diastolic blood pressure (DBP), non-fasting glucose (Glucose), total cholesterol, high density lipoprotein (HDL) cholesterol, low density lipoprotein (LDL) cholesterol and triglycerides), the most recent value (e.g. values measured in HUNT3) was used if available. If the individuals were not a part of HUNT3, measurements from HUNT2 were used. Age at participation was calculated to correspond with the health survey chosen. Blood pressure was taken three times during the clinical examination, and SBP and DBP measurements were calculated as the average of the second and third measurement. For individuals who only had two blood pressure measurements taken (12% of offspring in the mother-offspring pairs and 9% of offspring in the father-offspring pairs), the second measurement was used.

For the blood measurements, samples were taken from non-fasting participants. In HUNT3, participants’ total cholesterol was measured by enzymatic cholesterol esterase methodology; HDL cholesterol was measured by accelerator selective detergent methodology; triglycerides were measured by glycerol phosphate oxidase methodology; and glucose was measured by Hexokinase/G-6-PDH methodology (Abbott, Clinical Chemistry, USA). In HUNT2, participants’ total and HDL cholesterol and triglycerides were measured by applying enzymatic colorimetric cholesterol esterase methods (Boeheringer Mannheim, Mannheim, Germany) and glucose was measured by an enzymatic hexokinase method. The measurements are shown in millimole per liter. Weight and height were measured in light clothes and BMI was calculated as weight (kilograms) divided by the squared value of height (in metres).

We adjusted the blood pressure measurements of individuals who self-reported using blood pressure lowering medication by adding 15 mmHg to their SBP and 10 mmHg to their DBP. We chose this procedure over including medication use as a covariate to avoid introduction of possible collider biases into the analyses [32]. LDL cholesterol was calculated using the Friedewald formula [33]. All values more than 4 standard deviations from the mean were removed. If the variable was not normally distributed (triglycerides, BMI and non-fasting glucose) the values were natural log transformed before removing outlying values.

### Phenotypic relationship between birthweight and cardiometabolic risk factors

Own birthweight was available for individuals in HUNT after linking with the Medical Birth Registry of Norway (MBRN) [34]. The registry commenced in 1967, when health authorities began reporting pregnancy-related data; therefore, birthweight measurements were only available for HUNT participants born in 1967 or later. The validity of information on birthweight in the MBRN has previously been reported as very good [35]. To investigate if we could replicate the previously reported phenotypic associations between birthweight and cardiometabolic risk factors, we fit a linear mixed model to N=7,825 mother-offspring pairs and then N=6,875 father offspring pairs using the software package OpenMx [36] using the procedure described below. We modelled offspring cardiometabolic risk factor as the outcome and included offspring birthweight, offspring birthweight squared, offspring age, offspring sex and measurement occasion (HUNT2 or HUNT3) as fixed effects. The cryptic relatedness between offspring was modelled using a genetic relatedness matrix in the random effects part of the model as described below.

### Analysis of fetal growth and later life outcomes in the offspring

Cryptic relatedness is a problem for genetic studies of large population-based cohorts like HUNT. Whilst point estimates from genetic association analyses will often be unbiased in the presence of cryptic relatedness, standard errors can be too small, meaning that statistical tests of association may have inflated Type 1 error rates. Dropping one person from each pair of putatively related individuals is inefficient and requires an arbitrary threshold to be specified in order to declare a pair of individuals related (e.g. first order relatives). Thus, dropping individuals is unlikely to remove the non-independence of the error terms completely. In the case of single SNPs, this problem can be solved by using custom-written software packages. These software packages allow users to fit linear mixed models where a dataset is analysed as one large set of related individuals and the similarity between individuals is parameterized by a genome-wide genetic relationship matrix. However, these software packages are designed for GWAS analysis and may not have the flexibility to enable users to fit more complicated statistical models such as those involving genetic risk scores and conditional association analyses, as we wish to do here.

We therefore parameterized our statistical model using the OpenMx package [36] in the R statistics software. OpenMx allows users to model multivariate normal data flexibly in terms of fixed and random effects, and to estimate parameters simultaneously using full information maximum likelihood (FIML). We used the fixed effects part of the model to test for genetic association between the maternal GRS and offspring phenotype, and modelled the similarity between individuals in the random effects part of the model. The model for the fixed effects included terms for the genetic risk score of the mother (father), the genetic risk score of the offspring, age, sex and the measurement occasion (HUNT2 or HUNT3). In the random effects part of the model we modelled the similarity between individuals in terms of a genetic relationship matrix and an identity matrix for residual effects:

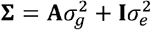

where Σ is the expected N × N phenotypic covariance matrix, **A** is an N × N genetic relationship matrix calculated using the GCTA software [37], **I** is an N × N identity matrix, 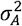 and 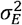 are variance components due to additive genetic and residual sources of variation respectively, and N is the number of individuals in the analysis.

Estimating these parameters using FIML in OpenMx is extremely computationally intensive in that it involves inverting a matrix of order N. Indeed our initial attempts to do this suggested that fitting the model to the HUNT data this way may not be possible given the limitations of our computing hardware. We therefore reparametrized the statistical model using a factor rotation which converted the problem from one involving an N × N matrix, to one involving a 1 × 1 matrix (see Supplementary Methods for details). Our implementation involved first performing a spectral decomposition of the genetic relationship matrix (**A**), and then pre-multiplying the matrices of outcomes and fixed effects respectively by the matrix of eigenvectors [38]. This pre-multiplication has the effect of “rotating away” the dependence between outcome trait values, leaving the random effects uncorrelated. The problem then reduces from N correlated observations (modelled by an N × N matrix), to N independent observations, greatly facilitating computation. An R script with code illustrating our method is included in the Supplementary Materials.

We first tested the relationship between maternal GRS and offspring birthweight in N=7,825 mother-offspring pairs and N=6,875 father offspring pairs to confirm that our GRS explained some of the variance in offspring birthweight. We then performed our primary analyses testing the relationship between maternal GRS and each of the offspring cardiovascular risk factors, whilst conditioning on offspring genotype at the same loci. We performed the same analyses in father-offspring pairs to assess whether there was evidence for a postnatal effect from either parent (Figure 1d). In addition to analysing all of the offspring together, we stratified the data into two groups; one group with offspring under age 40 at the time of measurement and one for offspring between 40 and 60 years of age. This was done to obtain a sample that would be easier to compare with the previous analysis of SBP in the UK Biobank Study by Warrington and colleagues [18]. Age strata for individuals over 60 is not presented due to the low number of individuals.

### Power calculations

We were interested in the statistical power of our approach to detect maternal genetic effects on offspring cardiometabolic risk factors. We therefore used the Maternal and Offspring Genetic Effects Power Calculator (https://evansgroup.di.uq.edu.au/MGPC/) to calculate power to detect association [39]. We assumed N = 26,057 complete mother offspring pairs, the absence of offspring genetic effects, and a Type 1 error rate of α = 0.05 (the presence/absence of offspring genetic effects has little influence on power to detect maternal genetic effects so long as the proportion of variance explained is small [39]).

## Results

### Phenotypic correlations

HUNT offspring with recorded values for birthweight were on average 31 years old, with a minimum age of 19, and a maximum age of 41 at the time of measurement used in this study. Descriptive statistics on the mother-offspring and father-offspring pairs are presented in Table 1. It’s important to note that only offspring born after 1967 had birthweight recorded and were included in this part of the analysis. Table 2 shows the phenotypic association between own birthweight and SBP, DBP, glucose, total, LDL and HDL cholesterol, triglycerides and BMI. Consistent with many previous observational epidemiological studies [40-43], linear regression yielded negative point estimates of the observational relationship between birthweight and blood pressure, LDL, total cholesterol and BMI. We also found evidence for positive quadratic terms in the model between birthweight and both BMI and glucose, suggesting U-shaped/J-shaped relationships between these variables. Finally, we found evidence for a positive linear relationship between HDL cholesterol and birthweight with additional evidence for a convex quadratic term indicating small and large babies are likely to have slightly reduced HDL levels in later life.

### Analysis of fetal growth and cardiometabolic risk factors in the HUNT offspring

We first checked whether the GRSs of birthweight associated SNPs from the latest GWAS of birthweight [18] were also related to offspring birthweight in HUNT. The full results are presented in Supplementary Table 3. In short, we found that maternal GRSs were strongly associated with increased offspring birthweight after conditioning on offspring GRS in HUNT. Offspring GRS was related to offspring birthweight, but this relationship attenuated after controlling for maternal GRS. In the case of the GRS consisting of SNPs that only had a maternal effect from the Warrington et al [18] birthweight GWAS, offspring GRS was not strongly related to offspring birthweight after controlling for maternal GRS. As expected, paternal GRS was not associated with offspring birthweight after conditioning on offspring GRS. The effect size of the offspring GRS was similar in mother-offspring and father-offspring pairs, and didn’t attenuate after adjusting for paternal GRS.

**Table 3:** Descriptive statistics for offspring cardiometabolic risk factors in the primary analyses

| Phenotype | Mother-Offspring pairs |  |  |  | Father-Offspring pairs |  |  |  |
| --- | --- | --- | --- | --- | --- | --- | --- | --- |
|  | N | Mean | SD | Range | N | Mean | SD | Range |
| Age | 26,057 | 41.4 | 12.7 | 19.1-83.2 | 19,792 | 39.3 | 12 | 19.1-84.8 |
| Sex (% male) | 26,057 | 48.4 | - | - | 19,792 | 48.4 | - | - |
| SBP (mmHg) | 25,946 | 128.3 | 17.3 | 70.0-218.0 | 19,711 | 126.9 | 16.4 | 70.0-218.0 |
| DBP (mmHg) | 25,940 | 73.9 | 11.9 | 36.0-134.0 | 19,711 | 72.8 | 11.5 | 38.0-126.0 |
| Glucose (mmol/L)* | 25,461 | 5.16 | 1.20 | 2.29-12.81 | 19,339 | 5.16 | 1.19 | 2.29-12.81 |
| Total Cholesterol (mmol/L) | 25,589 | 5.31 | 1.08 | 2.00-10.90 | 19,423 | 5.22 | 1.07 | 2.10-10.90 |
| LDL Cholesterol (mmol/L) | 25,533 | 3.26 | 0.95 | 0.14-8.60 | 19,392 | 3.19 | 0.93 | 0.27-8.60 |
| HDL Cholesterol (mmol/L) | 25,560 | 1.33 | 0.33 | 0.50-2.90 | 19,412 | 1.33 | 0.33 | 0.50-2.80 |
| Triglycerides (mmol/L)* | 25,916 | 1.35 | 1.73 | 0.18-11.70 | 19,680 | 1.32 | 1.73 | 0.49-11.70 |
| BMI* | 25,946 | 26.31 | 1.17 | 15.03-50.40 | 19,715 | 26.31 | 1.17 | 15.80-50.40 |

\*Offspring phenotype first (natural) logarithm transformed. SBP: systolic blood pressure; DBP: diastolic blood pressure; Glucose: non-fasting glucose; BMI: body mass index; LDL: low density lipoprotein; HDL: high density lipoprotein.

For the primary analyses investigating the effect of GRS on offspring cardiometabolic traits, we had a total of 26,057 mother-offspring pairs and 19,792 father-offspring pairs. HUNT offspring were on average 40 years old, with a minimum age of 19, and a maximum age of 85 at the time of measurement used in this study. Descriptive statistics on all of the outcome variables in the two samples are presented in Table 3. Our asymptotic power calculations indicated that we had (≥80%) power to detect a maternal genetic effect that explained as little as 0.04% of the variance in offspring outcome (N=26,057) (two tailed α = 0.05) and slightly lower power (>68%) (N=19,792) to detect a paternal genetic effect responsible for a similar proportion of the offspring phenotypic variance. Due to some missing data in the offspring’s cardiometabolic risk factors, the number of mother-offspring and father-offspring pairs differed slightly across the outcomes (Table 3). Although the sample size for some of the analyses are slightly lower (lowest being 25,461 mother-offspring pairs and 19,339 father-offspring pairs) we retain statistical power to detect an association of maternal GRS with offspring cardiometabolic risk factors (79% and 67% respectively) using the same parameters as above.

We found little evidence for an association between maternal (or paternal) GRS and any of the offspring cardiometabolic risk factors in later life, after adjusting for offspring GRS (Tables 4 and 5; Supplementary Table 4). These results hold for systolic blood pressure, which had previously been found to associate with maternal GRS in the Warrington et al GWAS of birthweight [18]. In contrast, there was strong evidence for a relationship between offspring GRS and some of the offspring phenotypes after conditioning on maternal GRS (Table 6). Specifically, there was evidence for a positive association between offspring GRS and both offspring glucose and LDL, and evidence for a negative relationship between offspring GRS and both systolic blood pressure and triglycerides.

**Table 4:** Results of regressing offspring cardiometabolic risk factors on maternal GRS* after conditioning on offspring GRS in mother-offspring pairs

| Outcome | Autosomal SNPs<br>(N=204) |  |  | Autosomal SNPs with maternal effect<br>(N=71) |  |  | Autosomal SNPs with maternal effect only<br>(N=31) |  |  |
| --- | --- | --- | --- | --- | --- | --- | --- | --- | --- |
|  | Effect<br>estimate | SE | p-value | Effect<br>estimate | SE | p-value | Effect<br>estimate | SE | p-value |
| SBP (mmHg) | -0.0083 | 0.0066 | 0.2135 | 0.0008 | 0.0066 | 0.8972 | -0.0056 | 0.0066 | 0.3968 |
| DBP (mmHg) | -0.0079 | 0.0065 | 0.2249 | -0.0049 | 0.0065 | 0.4512 | -0.0071 | 0.0064 | 0.2676 |
| Glucose (mmol/L)** | -0.0024 | 0.0071 | 0.7409 | -0.0021 | 0.0061 | 0.7854 | -0.0018 | 0.0070 | 0.8014 |
| Total Cholesterol (mmol/L) | -0.0019 | 0.0069 | 0.7810 | 0.0040 | 0.0068 | 0.5344 | -0.0034 | 0.0068 | 0.6185 |
| LDL Cholesterol (mmol/L) | -0.0045 | 0.0066 | 0.4938 | 0.0016 | 0.0066 | 0.8060 | -0.0016 | 0.0066 | 0.8091 |
| HDL Cholesterol (mmol/L) | 0.0045 | 0.0068 | 0.4993 | 0.0049 | 0.0065 | 0.4570 | 0.0095 | 0.0066 | 0.1527 |
| Triglycerides (mmol/L)** | 0.0016 | 0.0069 | 0.8148 | 0.0030 | 0.0069 | 0.6518 | -0.0097 | 0.0069 | 0.1628 |
| BMI** | -0.0127 | 0.0071 | 0.1631 | 0.0002 | 0.0068 | 0.8502 | -0.0118 | 0.0071 | 0.0910 |

\*Maternal and offspring GRS were coded so that increasing dosages reflected maternal alleles associated with increased offspring birthweight based on conditional GWAS results previously published.
\*\*Offspring phenotype first (natural) logarithm transformed. Effect estimates and standard errors are standardized. SBP: systolic blood pressure; DBP: diastolic blood pressure; Glucose: non-fasting glucose; BMI: body mass index; LDL: low density lipoprotein; HDL: high density lipoprotein. All analyses are adjusted for age, sex, participation period and GRS of offspring.

**Table 5:** Results of regressing offspring cardiometabolic risk factors on paternal GRS* after conditioning on offspring GRS in father-offspring pairs

| Outcome | Autosomal SNPs<br>(N=204) |  |  | Autosomal SNPs with maternal effect<br>(N=71) |  |  | Autosomal SNPs with maternal effect only<br>(N=31) |  |  |
| --- | --- | --- | --- | --- | --- | --- | --- | --- | --- |
|  | Effect estimate | SE | p-value | Effect estimate | SE | p-value | Effect estimate | SE | p-value |
| SBP (mmHg) | -0.0066 | 0.0077 | 0.3871 | -0.0008 | 0.0076 | 0.9143 | 0.0097 | 0.0076 | 0.2050 |
| DBP (mmHg) | -0.0054 | 0.0075 | 0.4729 | 0.0061 | 0.0074 | 0.4114 | 0.0025 | 0.0075 | 0.7412 |
| Glucose (mmol/L)** | 0.0055 | 0.0081 | 0.4685 | 0.0086 | 0.0080 | 0.2786 | 0.0017 | 0.0075 | 0.8362 |
| Total Cholesterol (mmol/L) | -0.0019 | 0.0079 | 0.8058 | -0.0085 | 0.0078 | 0.2798 | -0.0102 | 0.0079 | 0.1951 |
| LDL Cholesterol (mmol/L) | -0.0009 | 0.0075 | 0.9056 | -0.0019 | 0.0074 | 0.8027 | -0.0033 | 0.0075 | 0.6637 |
| HDL Cholesterol (mmol/L) | -0.0002 | 0.0078 | 0.8380 | -0.0046 | 0.0078 | 0.5494 | -0.0131 | 0.0077 | 0.0959 |
| Triglycerides (mmol/L)** | $1.35 \times 10^{-05}$ | 0.0080 | 0.9955 | -0.0039 | 0.0079 | 0.6087 | -0.0039 | 0.0079 | 0.6238 |
| BMI** | 0.0018 | 0.0082 | 0.8205 | -0.0001 | 0.0068 | 1 | 0.0119 | 0.0080 | 0.1414 |

\*Paternal and offspring GRS were coded so that increasing dosages reflected maternal alleles associated with increased offspring birthweight based on conditional GWAS results previously published.
\*Offspring phenotype first (natural) logarithm transformed. Effect estimates and standard errors are standardized. SBP: systolic blood pressure; DBP: diastolic blood pressure; Glucose: non-fasting glucose; BMI: body mass index; Glucose: non-fasting glucose; LDL: low density lipoprotein; HDL: high density lipoprotein. All analyses are adjusted for age, sex, participation period and GRS of offspring.

**Table 6:** Results of regressing offspring cardiometabolic risk factors on offspring GRS* after conditioning on maternal GRS in mother-offspring pairs

| Outcome | Autosomal SNPs<br>(N=204) |  |  | Autosomal SNPs with maternal effect<br>(N=71) |  |  | Autosomal SNPs with maternal effect only<br>(N=31) |  |  |
| --- | --- | --- | --- | --- | --- | --- | --- | --- | --- |
|  | Effect<br>estimate | SE | p-value | Effect<br>estimate | SE | p-value | Effect<br>estimate | SE | p-value |
| SBP (mmHg) | -0.0105 | 0.0065 | 0.1093 | **-0.0170** | **0.0065** | **0.0089** | **-0.0214** | **0.0066** | **0.0011** |
| DBP (mmHg) | 0.0027 | 0.0064 | 0.6656 | -0.0047 | 0.0063 | 0.4644 | -0.0114 | 0.0063 | 0.0733 |
| Glucose (mmol/L)** | **0.0229** | **0.0070** | **0.0007** | **0.0247** | **0.0059** | **2.69x10<sup>-5</sup>** | **0.0180** | **0.0068** | **0.0082** |
| Total Cholesterol (mmol/L) | 0.0068 | 0.0067 | 0.2951 | 0.0083 | 0.0060 | 0.1594 | 0.0128 | 0.0067 | 0.0561 |
| LDL Cholesterol (mmol/L) | 0.0090 | 0.0064 | 0.1479 | **0.0141** | **0.0064** | **0.0271** | **0.0154** | **0.0064** | **0.0162** |
| HDL Cholesterol (mmol/L) | 0.0015 | 0.0066 | 0.8152 | 0.0049 | 0.0065 | 0.3956 | 0.0072 | 0.0066 | 0.2680 |
| Triglycerides (mmol/L)** | 0.0117 | 0.0068 | 0.0866 | **-0.0218** | **0.0068** | **0.0012** | **-0.0133** | **0.0068** | **0.0494** |
| BMI** | -0.0022 | 0.0069 | 0.7783 | -0.0068 | 0.0068 | 0.3381 | -0.0099 | 0.0069 | 0.1320 |

\*Maternal and offspring GRS were coded so that increasing dosages reflected maternal alleles associated with increased offspring birthweight based on conditional GWAS results previously published.
\*Offspring phenotype first (natural) logarithm transformed. Effect estimates and standard errors are standardized. SBP: systolic blood pressure; DBP: diastolic blood pressure; Glucose: non-fasting glucose; BMI: body mass index; LDL: low density lipoprotein; HDL: high density lipoprotein. All analyses are adjusted for age, sex, participation period and GRS of offspring. Results with p-values less than 0.05 are shown in bold.

#### Age stratified analyses

Cardiometabolic pathology becomes more apparent with increasing age. Indeed, it is possible that younger individuals within the HUNT Study do not show observable compensatory changes in cardiometabolic risk factors, reducing the power of our analyses to detect evidence for the observational associations between birthweight and cardiometabolic risk factors to be causal. We therefore divided our dataset into two strata based on age of the offspring (i.e. offspring under 40 years of age and offspring between 40 and 60 years of age). Our asymptotic power calculations indicated that we had (≥80%) power to detect a maternal genetic effect that explained as little as 0.09% of the variance in offspring SBP (N=12,037 and N=11,849) (α = 0.05) and slightly lower power (>66%) (N=10,393 and N=8,402) to detect a paternal genetic effect responsible for a similar proportion of the offspring phenotypic variance. Table 7 shows the main results of the stratified analyses compared with those previously reported in the UK BioBank by Warrington et al in their GWAS of birthweight [18]. Whereas Warrington and colleagues found a significant positive effect of maternal GRS on offspring SBP when adjusting for offspring GRS, we find no effect in the stratified analyses.

**Table 7:** Association between maternal SNPs influencing offspring birthweight and offspring SBP after conditioning on offspring genotype in different age strata compared with previous results from Warrington et al [18].

|  |  | Autosomal SNPs<br>(N=204) |  |  | Autosomal SNPs with maternal effect<br>(N=71) |  |  | Autosomal SNPs with maternal effect<br>only (N=31) |  |  |
| --- | --- | --- | --- | --- | --- | --- | --- | --- | --- | --- |
| Analysis sample | N | Effect<br>estimate | SE | p-value | Effect<br>estimate | SE | p-value | Effect<br>estimate | SE | p-value |
| Age 20-40 in HUNT: |  |  |  |  |  |  |  |  |  |  |
| Mother-Offspring pairs | 12,037 | -0.015 | 0.0095 | 0.11 | -0.0046 | 0.0095 | 0.63 | -0.0092 | 0.0095 | 0.33 |
| Father-Offspring pairs | 10,393 | -0.0059 | 0.010 | 0.56 | -0.013 | 0.010 | 0.21 | -0.0025 | 0.010 | 0.81 |
| Age 40-60 in HUNT: |  |  |  |  |  |  |  |  |  |  |
| Mother-Offspring pairs | 11,849 | -0.0048 | 0.010 | 0.65 | -0.0061 | 0.0087 | 0.65 | 0.0011 | 0.010 | 0.91 |
| Father-Offspring pairs | 8,402 | -0.0067 | 0.012 | 0.59 | 0.0085 | 0.012 | 0.47 | 0.018 | 0.012 | 0.15 |
| UK Biobank results from<br>Warrington<br>et al [18]: |  |  |  |  |  |  |  |  |  |  |
| Mother-Offspring pairs** | 3,886 | 0.043 | 0.030 | 0.15 | **0.12** | **0.050** | **0.018** | **0.21** | **0.083** | **0.011** |
| Father-Offspring pairs** | 1,749 | 0.032 | 0.044 | 0.46 | 0.091 | 0.075 | 0.221 | 0.029 | 0.13 | 0.82 |

All analysis are adjusted for age, sex, participation period and GRS of offspring. Results with p-values less than 0.05 are shown in bold. Effect estimates and standard errors in HUNT are standardized.
\*\*Maternal, paternal and offspring GRS were coded so that increasing dosages reflected maternal alleles associated with increased offspring birthweight based on conditional GWAS results previously published.
\*\*Unstandardized values.

## Discussion

The Developmental Origins of Health and Disease (DOHaD) hypothesis posits that adverse environmental factors *in utero* or in the early years of life result in increased future risk of cardiometabolic disease [1, 5, 7]. In this study, we used a Mendelian randomization paradigm to provide evidence for or against the existence of DOHaD mechanisms that are related to fetal growth and lower birth weight for a range of cardiometabolic risk factors [16, 18]. Specifically, we tested whether a genetic risk score in mothers intended to proxy for maternal intrauterine influences on offspring birthweight was also associated with offspring cardiometabolic risk factors, whilst simultaneously conditioning on offspring GRS constructed from the same birthweight associated loci. There was no strong evidence of association in a sample of over 25,000 mother-offspring pairs from the Norwegian HUNT study, implying that if such an effect on cardiometabolic risk factors exists, it may be small compared to other sources of inter-individual variation, or only affects a few individuals.

Our study is the largest parent-offspring Mendelian randomization study of DOHaD performed to date. The HUNT Study contains over 25,000 genotyped mother-offspring pairs where the majority of the offspring are middle-aged adults, and are therefore old enough to have begun developing observable signs of cardiometabolic disease. Our asymptotic calculations indicated that we had strong (≥80%) power to detect a maternal genetic effect that explained as little as 0.04% of the variance in offspring outcome (two tailed α= 0.05). In contrast, our previous study in the UK Biobank [18] (where we first used this Mendelian randomization paradigm to investigate DOHaD), involved only 3,886 mother-offspring pairs, and was likely underpowered. Interestingly, Warrington and colleagues found evidence for a positive relationship between maternal birthweight lowering SNPs and reduced offspring SBP (i.e. the opposite of what DOHaD would predict), however this result did not replicate in our sample. Possible reasons for the discrepancy include the differences in sample ascertainment across the studies, or that the younger offspring in HUNT did not manifest a large enough effect [18]. When stratifying our analysis by age, we did find effects in the same direction as our original study for the 40-60 years age group; however, the statistical support for the effect was extremely weak. Taken together, the UK BioBank and HUNT results provide converging evidence that maternal genetic effects that predispose to low offspring birthweight are *not* associated with *increased* systolic blood pressure in later life.

In contrast, we did find evidence for association between offspring GRS and a number of offspring cardiometabolic risk factors, even after conditioning on maternal GRS. These results are broadly consistent with the Fetal Insulin hypothesis [44-47] and previous studies that have used LD score regression and G-REML approaches to suggest that much of the phenotypic correlation between birthweight and cardiometabolic risk is driven by genetic pleiotropy in the offspring genome rather than DOHAD mechanisms [18, 48]. We note that the direction of the associations involving the offspring GRS and offspring phenotypes are a little difficult to interpret, since the GRS were defined on the basis of maternal genotypic effects on offspring birthweight, whereas these reported associations involve offspring GRS. Offspring genotypes at some of the same loci are known to have quantitatively and qualitatively different effects on offspring birthweight (including the direction of association) compared to the maternal effects. Nevertheless our results show clearly that maternal SNPs that influence offspring birthweight have pleiotropic effects on offspring cardiometabolic traits when these same SNPs are transmitted to their offspring.

Another novel facet of our study was the use of the OpenMx software package to model the complicated data structure within the HUNT Study. Using traditional formulations of FIML to model the relatedness structure using a genetic relationship matrix would be computationally prohibitive with the HUNT sample, as maximizing the likelihood would involve an expensive inversion of a matrix of order N. In contrast, our implementation permits complicated tests of association to be performed in the fixed effects part of the model, whilst simultaneously modelling cryptic relatedness in the random effects part of the model in a computationally efficient manner [38]. We hope that our implementation will prove useful in complicated genetic analyses of other large scale population-based cohorts where cryptic relatedness/population stratification is likely to be an issue. We have included an example R script in the Supplementary Materials section of the manuscript that can be used as a template by interested researchers. We caution users, however, that specification of the covariance part of the model is more rigid using our speed up in that only two variance components can be fitted simultaneously, one being a residual variance component that is uncorrelated across individuals.

Our approach has a number of limitations which we discuss in the remaining paragraphs. First, we assume that the maternal SNPs that affect offspring birthweight do so via fetal growth (as reflected in birthweight). This is important, because as many others have noted, it may not be fetal growth/birthweight itself that is relevant for the validity of DOHaD. Rather it could be poor development of different key organs, in key stages of the pregnancy or a particular adverse maternal environment due to famine, disease or a range of other factors. Indeed, it would likely be profitable to use the same framework to investigate the association between offspring cardiometabolic disease and other adverse maternal exposures, such as maternal BMI, maternal alcohol consumption, preeclampsia and gestational diabetes. Their effect may be qualitatively and quantitatively different from the maternal effect on birthweight within healthy subjects delivering babies within the normal range. However, even though the mechanisms through which our maternal SNPs influence offspring birthweight are largely unknown, we know that they play an important part in fetal growth of the offspring. Further Mendelian randomization studies on different maternal exposures are warranted including on those that do not necessarily exert observable effects on offspring birthweight.

Second, our example here, and Mendelian randomization approaches in general, typically test small changes in an exposure. However, it may be that DOHaD mechanisms are important in the genesis of cardiometabolic risk, but only in the case of severe exposures (e.g. famine or obesity) at the extreme ends of the spectrum. These effects may be qualitatively different from small perturbations in the environment that produce relatively subtle variations in the normal healthy population. If DOHaD is only relevant in the case of extreme environmental effects, then Mendelian randomization approaches applied to population data may not be well suited to testing the hypothesis.

Third, although our methods rely on Mendelian randomization principles to inform on the validity of DOHaD (i.e. we use genetic variants to increase our study’s robustness to environmental confounding), we did not perform formal instrumental variables analyses in this manuscript. The reason is that we do not have appropriate estimates of the effect of maternal genotypes on the intrauterine environment. We only have estimates of the relationship between SNPs and offspring birthweight, which is an imperfect proxy of fetal growth restriction. Therefore, it does not make sense to estimate causal effect sizes in our study as in typical MR analyses. However, we note that it may be possible to estimate the effect of a putative latent variable indexing growth restriction using, for example, latent variable models; this is an area of future research for our group.

Fourth, our power calculations show that we were well powered (>80% at α = 0.05) to detect an association between maternal genetic risk score and offspring cardiometabolic risk factors responsible for as little as 0.04% of the phenotypic variance. However, whilst our study is the largest and most powerful genetic investigation into DOHaD to date, the actual variance in the offspring cardiometabolic risk factor explained by the maternal GRS, depends critically upon the underlying genetic model, and could be even smaller than 0.04%. In an attempt to make this clear, Figure 2 is a path diagram that illustrates the relationship between maternal GRS, offspring GRS, an intrauterine environment that reduces fetal growth (modelled as a single latent unobserved variable), offspring birthweight and an offspring cardiometabolic risk factor. In this diagram, and consistent with most formulations of DOHaD, we assume that (i) there is no direct causal effect of birthweight on cardiometabolic risk (i.e. no arrow from birthweight to the cardiometabolic risk factor), and (ii) no effect of maternal GRS on the offspring cardiometabolic risk factor that goes through paths other than fetal growth restriction (e.g. no post-natal mechanisms). To make calculations and explication easier, we assume that all variables have been standardized to unit variance. Under this model, the correlation between birthweight and the cardiometabolic risk factor is a function of two processes. One is the effect of the intrauterine environment on birthweight and the cardiometabolic risk factors (i.e. the product of path coefficients λ_1_ and λ_2_). The second is the residual covariance between birthweight and the cardiometabolic risk factors. This latter pathway includes both environmental factors other than fetal growth restriction that affect both phenotypes and the effect of polygenes that are not modelled in the experiment whose joint effects are quantified by the parameter Θ. These correlations could be positive or negative individually, but when combined produce a very small (|r| <= 0.05) negative phenotypic correlation between birthweight and most of the cardiometabolic risk factors. The point is that, unless the residual covariance between birthweight and the cardiometabolic risk factor is positive, the values for path coefficients λ_1_ and λ_2_ are likely to be very small in order to be consistent with the observed phenotypic correlations.

**Figure 2:**
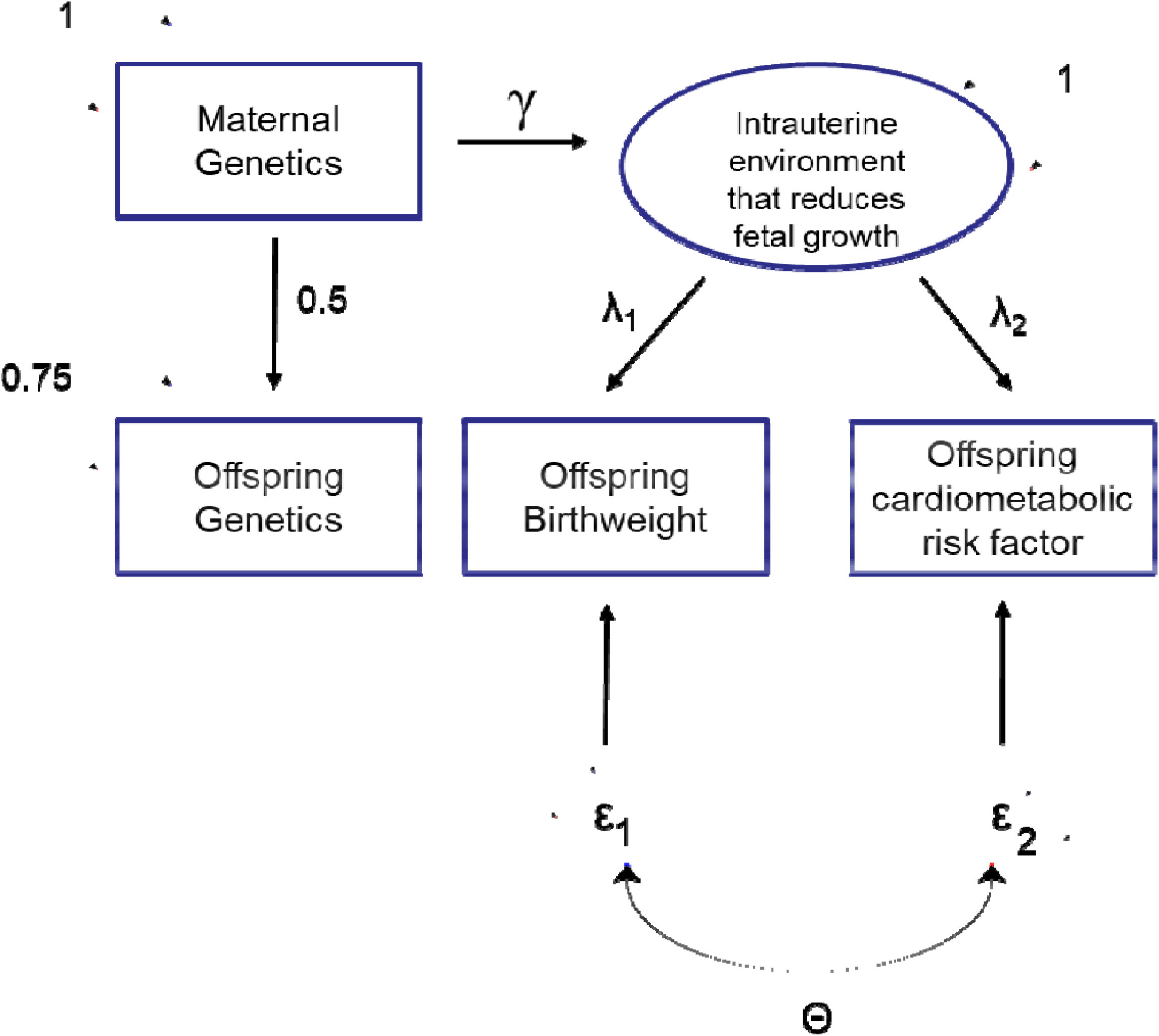
Path diagram of the relationship between maternal GRS, offspring GRS, the intrauterine environment, offspring birthweight and an offspring cardiometabolic risk factor. Variables within square boxes represent observed variables, whereas variables in circles represent latent unobserved variables. Unidirectional arrows represent causal relationships from tail to head, whilst two headed arrows represent correlational relationships. Greek letters on one headed arrows represent path coefficients which quantify the expected causal effect of one variable on the other. Greek letters on two headed arrows represent covariances between variables. The two epsilon variables represent residual latent factors (both environmental and genetic) that are not modelled in the study. The coefficient Θ represents the covariance between the residual terms. We assume that all variables are standardized to have unit variance. Consequently, the residual variance of the offspring GRS is set to 0.75 since ¼ of the variance comes from the maternal genotype. For the purposes of the power calculation described in the discussion, we assume that maternal SNPs that affect offspring birthweight do so through a single latent intrauterine factor, and that this factor also exerts long term effects on the offspring cardiometabolic risk factor of interest.

The variance in birthweight explained by the maternal GRS is a function of the direct association between the SNPs and the intrauterine environment (the path coefficient γ), and the effect of the intrauterine environment on birthweight (the path coefficient λ_1_ - the precise formula being: γ^2^λ_1_ ^2^).The variance explained in the cardiometabolic risk factor by the maternal GRS is equal to the product of the SNPs’ direct effect on the intrauterine environment (path coefficient γ in Figure 2), multiplied by the effect of the intrauterine environment on the cardiometabolic risk factor (path coefficient λ_2_ in Figure 2) all squared. There are an infinite number of ways these parameters can vary to make the underlying model consistent with the pattern of observed correlations and the proportion of variance explained in birthweight by the maternal GRS. To give the reader an idea of the potentially small numbers involved, we assume that the correlation between birthweight and the cardiometabolic risk factor is completely explained by the intrauterine environment and λ_1_ = −0.5 and λ_2_ = 0.1 (so that the observed correlation r = λ_1_ λ_2_ = −0.05). In order for the underlying model to also be consistent with the maternal GRS explaining a small percentage of the variance in birthweight (say 0.5% of the variance), then the path coefficient between the maternal GRS and the latent intrauterine variable γ would equal 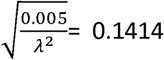. These values in turn would imply that the variance explained in the cardiometabolic risk factor by the maternal genetic risk score would be 0.1414^2^ × 0.1^2^ = 0.02%, which is a small proportion of the variance, and one that we are only moderately well powered to detect (>50%) in our study. Our point, however, is that the proportion of variance in the outcome explained by the maternal GRS may be very small, and so power may only be moderate despite the very large sample size of HUNT. The corollary to this though, is that we are very well powered to detect larger effects of the intrauterine environment influencing offspring birthweight on cardiometabolic risk factors, and the fact that we do not detect these suggests that if such an effect is present, it is likely to be small.

Finally, we recognize that our act of conditioning on offspring GRS, may have induced a (spurious) correlation between maternal GRS and paternal GRS due to conditioning on a collider variable, potentially biasing the results of our maternal GRS analyses. However, any such bias is likely to be small in magnitude as it relies on the existence of (and is proportional to the size of) direct paternal genetic effects from the same SNPs on the offspring phenotype. As sizeable paternal genetic effects on offspring cardiometabolic risk are unlikely at these loci, we doubt that collider bias is a serious impediment to the validity of our study [49].

In conclusion, we did not find evidence for a causal effect of the intrauterine environment (as proxied by maternal genetic effects on offspring birthweight) on offspring cardiometabolic risk factors in a population-based sample of individuals. We did, however, find evidence of genetic pleiotropy between offspring birthweight and offspring cardiometabolic risk factors which helps explain the robust observational relationships between the variables.

## Data Availability

Researchers with a PhD associated with Norwegian research institutes can apply for the use of HUNT material: data and samples - given approval by a Regional Committee for Medical and Health Research Ethics. Researchers from other countries are welcome to apply in cooperation with a Norwegian Principle Investigator. https://www.ntnu.edu/hunt/data

## Acknowledgments

We would like to thank the research participants of the HUNT study. The Nord-Trøndelag Health Study (The HUNT Study) is a collaboration between HUNT Research Centre (Faculty of Medicine and Health Sciences, NTNU, Norwegian University of Science and Technology), Nord-Trøndelag County Council, Central Norway Regional Health Authority, and the Norwegian Institute of Public Health. The genotyping in HUNT was financed by the National Institutes of Health (NIH); University of Michigan; The Research Council of Norway; The Liaison Committee for Education, Research and Innovation in Central Norway; and the Joint Research Committee between St. Olavs hospital and the Faculty of Medicine and Health Sciences, NTNU. The genotype quality control and imputation has been conducted by the K.G. Jebsen Center for Genetic Epidemiology, Department of Public Health and Nursing, Faculty of Medicine and Health Sciences, NTNU. This research was carried out at the Translational Research Institute, Woolloongabba, QLD 4102, Australia. The Translational Research Institute is supported by a grant from the Australian Government. Support for this research have been given by the Norwegian Diabetes Association and Nils Normans minnegave. G.H.M is supported by the Norwegian Research Council (Post doctorial mobility research grant 287198). N.M.W. is supported by an Australian National Health and Medical Research Council Early Career Fellowship (APP1104818). MCN and OpenMx development were funded by NIH grant DA-018673. D.A.L is supported by the British Heart Foundation (AA/18/7/34219) and European Research Council (669545). D.A.L, G.D.S, D.M.E are affiliated with a Unit that receives support from the University of Bristol and the UK Medical Research Council (MC_UU_00011/1 and MC_UU_00011/6). D.M.E. is funded by an Australian National Health and Medical Research Council Senior Research Fellowship (APP1137714) and NHMRC project grants (GNT1125200, GNT1157714, GNT1183074).

## Notes

### Competing Interest Statement

D.A.L. has received research support from several
national and international government and charity funders, and from Medtronic Ltd and Roche Diagnostics in the last 10 years.

## References

1. Barker, D.J., The fetal and infant origins of adult disease. Bmj, 1990. 301(6761): p. 1111.

2. Forsdahl, A., Are poor living conditions in childhood and adolescence an important risk factor for arteriosclerotic heart disease? Br J Prev Soc Med, 1977. 31(2): p. 91–5.

3. Hales, C.N. and D.J. Barker, Type 2 (non-insulin-dependent) diabetes mellitus: the thrifty phenotype hypothesis. Diabetologia, 1992. 35(7): p. 595–601.

4. Roseboom, T.J., et al., Effects of prenatal exposure to the Dutch famine on adult disease in later life: an overview. Mol Cell Endocrinol, 2001. 185(1-2): p. 93–8.

5. Gillman, M.W., Developmental origins of health and disease. N Engl J Med, 2005. 353(17): p. 1848–50.

6. Godfrey, K.M. and D.J. Barker, Fetal nutrition and adult disease. The American Journal of Clinical Nutrition, 2000. 71(5): p. 1344s–1352s.

7. Barker, D.J., et al., Fetal nutrition and cardiovascular disease in adult life. Lancet, 1993. 341(8850): p. 938–41.

8. Seghieri, G., et al., Relationship between gestational diabetes mellitus and low maternal birth weight. Diabetes Care, 2002. 25(10): p. 1761–5.

9. Dickinson, H., et al., A review of fundamental principles for animal models of DOHaD research: an Australian perspective. J Dev Orig Health Dis, 2016. 7(5): p. 449–472.

10. Smith, G.D. and S. Ebrahim, ‘Mendelian randomization’: can genetic epidemiology contribute to understanding environmental determinants of disease? Int J Epidemiol, 2003. 32(1): p. 1–22.

11. Huang, T., et al., Association of Birth Weight With Type 2 Diabetes and Glycemic Traits: A Mendelian Randomization Study. JAMA Netw Open, 2019. 2(9): p. e1910915.

12. Wang, T., et al., Low birthweight and risk of type 2 diabetes: a Mendelian randomisation study. Diabetologia, 2016. 59(9): p. 1920–7.

13. Zanetti, D., et al., Birthweight, Type 2 Diabetes Mellitus, and Cardiovascular Disease: Addressing the Barker Hypothesis With Mendelian Randomization. Circ Genom Precis Med, 2018. 11(6): p. e002054.

14. Shannon D’Urso, G.W., Liang-Dar Hwang, Gunn-Helen Moen, Nicole M Warrington, David M Evans, A Cautionary Note on Using Mendelian Randomization to Examine the Developmental Origins of Health and Disease (DOHaD) Hypothesis. Submitted: Journal of Developmental Origins of Health and Disease.

15. Brumpton, B., et al., Within-family studies for Mendelian randomization: avoiding dynastic, assortative mating, and population stratification biases. bioRxiv, 2019: p. 602516.

16. Evans, D.M., et al., Elucidating the role of maternal environmental exposures on offspring health and disease using two-sample Mendelian randomization. Int J Epidemiol, 2019. 48(3): p. 861–875.

17. Didelez, V. and N. Sheehan, Mendelian randomization as an instrumental variable approach to causal inference. Stat Methods Med Res, 2007. 16(4): p. 309–30.

18. Warrington, N.M., et al., Maternal and fetal genetic effects on birth weight and their relevance to cardio-metabolic risk factors. Nat Genet, 2019. 51(5): p. 804–814.

19. Richmond, R.C., et al., Using Genetic Variation to Explore the Causal Effect of Maternal Pregnancy Adiposity on Future Offspring Adiposity: A Mendelian Randomisation Study. PLOS Medicine, 2017. 14(1): p. e1002221.

20. Freathy, R.M., Can genetic evidence help us to understand the fetal origins of type 2 diabetes? Diabetologia, 2016. 59(9): p. 1850–4.

21. Krokstad, S., et al., Cohort Profile: the HUNT Study, Norway. Int J Epidemiol, 2013. 42(4): p. 968–77.

22. Holmen, T.L., et al., Cohort profile of the Young-HUNT Study, Norway: A population-based study of adolescents. International Journal of Epidemiology, 2013. 43(2): p. 536–544.

23. Ferreira, M.A., et al., Shared genetic origin of asthma, hay fever and eczema elucidates allergic disease biology. Nat Genet, 2017. 49(12): p. 1752–1757.

24. Dunham, I., et al., An integrated encyclopedia of DNA elements in the human genome. Nature, 2012. 489(7414): p. 57–74.

25. Wang, C., et al., Ancestry estimation and control of population stratification for sequence-based association studies. Nat Genet, 2014. 46(4): p. 409–15.

26. Li, J.Z., et al., Worldwide human relationships inferred from genome-wide patterns of variation. Science, 2008. 319(5866): p. 1100–4.

27. Chang, C.C., et al., Second-generation PLINK: rising to the challenge of larger and richer datasets. Gigascience, 2015. 4: p. 7.

28. Loh, P.R., et al., Reference-based phasing using the Haplotype Reference Consortium panel. Nat Genet, 2016. 48(11): p. 1443–1448.

29. Das, S., et al., Next-generation genotype imputation service and methods. Nat Genet, 2016. 30.

30. McCarthy, S., et al., A reference panel of 64,976 haplotypes for genotype imputation. Nat Genet, 2016. 48(10): p. 1279–83.

31. Manichaikul, A., et al., Robust relationship inference in genome-wide association studies. Bioinformatics, 2010. 26(22): p. 2867–2873.

32. Tobin, M.D., et al., Adjusting for treatment effects in studies of quantitative traits: antihypertensive therapy and systolic blood pressure. Stat Med, 2005. 24(19): p. 2911–35.

33. Friedewald, W.T., R.I. Levy, and D.S. Fredrickson, Estimation of the Concentration of Low-Density Lipoprotein Cholesterol in Plasma, Without Use of the Preparative Ultracentrifuge. Clinical Chemistry, 1972. 18(6): p. 499–502.

34. Norwegian Institute of Public Health. Medical Birth Registry of Norway Available at: www.fhi.no/en/hn/health-registries/medical-birth-registry-of-norway/medical-birth-registry-of-norway. [cited 2019].

35. Moth, F.N., et al., Validity of a selection of pregnancy complications in the Medical Birth Registry of Norway. Acta Obstet Gynecol Scand, 2016. 95(5): p. 519–27.

36. Boker, S., et al., OpenMx: An Open Source Extended Structural Equation Modeling Framework. Psychometrika, 2011. 76(2): p. 306–317.

37. Yang, J., et al., GCTA: a tool for genome-wide complex trait analysis. Am J Hum Genet, 2011. 88(1): p. 76–82.

38. Meyer, K., Maximum Likelihood Estimation of Variance Components for a Multivariate Mixed Model with Equal Design Matrices. Biometrics, 1985. 41(1): p. 153–165.

39. Moen, G.-H., et al., Calculating Power to Detect Maternal and Offspring Genetic Effects in Genetic Association Studies. Behavior Genetics, 2019. 49(3): p. 327–339.

40. Wurtz, P., et al., Metabolic signatures of birthweight in 18 288 adolescents and adults. Int J Epidemiol, 2016. 45(5): p. 1539–1550.

41. Huxley, R.R., A.W. Shiell, and C.M. Law, The role of size at birth and postnatal catch-up growth in determining systolic blood pressure: a systematic review of the literature. J Hypertens, 2000. 18(7): p. 815–31.

42. Lawlor, D.A., et al., Sex differences in the association between birth weight and total cholesterol. A meta-analysis. Ann Epidemiol, 2006. 16(1): p. 19–25.

43. Newsome, C.A., et al., Is birth weight related to later glucose and insulin metabolism?--A systematic review. Diabet Med, 2003. 20(5): p. 339–48.

44. Hattersley, A.T. and J.E. Tooke, The fetal insulin hypothesis: an alternative explanation of the association of low birthweight with diabetes and vascular disease. Lancet, 1999. 353(9166): p. 1789–92.

45. Hattersley, A.T., et al., Mutations in the glucokinase gene of the fetus result in reduced birth weight. Nature Genetics, 1998. 19(3): p. 268–270.

46. Freathy, R.M., et al., Type 2 diabetes risk alleles are associated with reduced size at birth. Diabetes, 2009. 58(6): p. 1428–33.

47. Tyrrell, J.S., et al., Parental diabetes and birthweight in 236 030 individuals in the UK biobank study. Int J Epidemiol, 2013. 42(6): p. 1714–23.

48. Horikoshi, M., et al., Genome-wide associations for birth weight and correlations with adult disease. Nature, 2016. 538(7624): p. 248–252.

49. Lawlor, D., et al., Using Mendelian randomization to determine causal effects of maternal pregnancy (intrauterine) exposures on offspring outcomes: Sources of bias and methods for assessing them. Wellcome Open Res, 2017. 2: p. 11.

